# Impact of Residential Neighborhood and Race/Ethnicity on Outcomes of Hospitalized Patients with COVID-19 in the Bronx

**DOI:** 10.1101/2021.01.09.21249515

**Authors:** Dwayvania Miller, Amara Sarwal, Bo Yu, Edgar Gomez, Victor Perez-Gutierrez, Marcia Gossai, Elisenda Valdez, Astrid Mendez, Sarah Chaudry, Usha Venugopal, Vihren Dimitrov, Moiz Kasubhai, Vidya Menon

## Abstract

The socially vulnerable have been most affected due to the COVID-19 pandemic, similar to the aftermath of any major disaster. Racial and social minorities are experiencing a disproportionate burden of morbidity and mortality.

The aim of this study was to evaluate the impact of residential location/community and race/ethnicity on outcomes of COVID-19 infection among hospitalized patients within the Bronx. This was a single center retrospective observational cohort study that included SARS-CoV2 positive adult residents of the Bronx (stratified as residents of South Bronx vs Rest of Bronx) hospitalized between March-May 2020. Data extracted from hospital electronic medical records included residential addresses, race, comorbidities, and insurance details. Comorbidity burden other clinical and laboratory details were also assessed to determine their correlation to COVID-19 severity of illness and outcomes of mortality and length of stay.

As expected, the COVID-19 pandemic differentially affected outcomes in those in the more socially disadvantaged area of the South Bronx versus the rest of the Bronx borough. Residents of the South Bronx had a significantly higher comorbidity burden and had public insurance to access medical care in comparison to the remainder of the Bronx. Interestingly, for the patient population studied there was no observed difference in 30-day mortality by race/ethnicity among those infected with COVID- 19 in spite of the increased disease burden observed.

This adds an interesting perspective to the current literature, and highlights the need to address the social/economic factors contributing to health access disparity to reduce the adverse impact of COVID-19 in these communities.

## Introduction

The socially vulnerable are at highest risk of adverse outcomes following any disaster^2,3^. Racial and ethnic minorities are experiencing a disproportionate burden of morbidity and mortality due to COVID-19 infection^4^. Existing socioeconomic disparities affect access to healthcare across NYC including response and outcomes of the infection. As of April 30, 2020, local NYC data showed that there were 164,505 cases of COVID-19 confirmed.^1^ Case allocation per borough found Queens to have 51,028 cases (31%); Brooklyn with 43,621 cases (27%); Bronx with 37,551 cases (23%); Manhattan with 20,363 cases (12%) and Staten Island with 11,861 cases (7%).^9^ It is to be noted that the 3 boroughs most affected also have the lowest 3 median household incomes in NYC with the Bronx being the poorest. As it is related to ethnicity, COVID- 19 related fatality was 34% among Hispanics, 28% in Blacks, 27% among Whites, and 7% among Asians in NYC^6^.

Our facility is a city hospital that provides healthcare primarily to the South Bronx, where approximately 29% of the population live in poverty with high unemployment rates, 14% of adults are uninsured and 10% have foregone medical care in the past 12 months.^6^ Our facility treated the highest amount of SARS-CoV2 positive patients within the South Bronx, and as of April 2020, LMC had served approximately 1,075 patients that tested positive for COVID-19. The aim of our study was to evaluate the impact of residential location/community and race/ethnicity on outcomes of COVID-19 infection among hospitalized patients.

## Methods

This was a single center retrospective observational cohort study that included SARS-CoV2 positive Bronx adult residents hospitalized between March to May 2020. Pregnant patients & those living outside the Bronx were excluded. Data was collected from the hospital electronic medical records. Residential address based on zip code was obtained to create two cohorts (South Bronx (SB) and rest of the Bronx (RB)^9^. Comorbidity burden, assessed using the Charlson Comorbidity Index, as well as insurance status were evaluated in comparison to COVID-19 severity of illness and outcome. Outcomes were mortality and length of stay (LOS). Descriptive statistics of variables were summarized as median and 25-75 IQR. Chi-Square/Fisher-Exact tests for categorical variables, Mann-Whitney U test for numeric variables & multiple logistic regression to evaluate the covariates to mortality was used. p <0.05 was considered significant. IBM SPSS v22 for Windows was used for analysis.

## Results

Of the 977 patients included in study, 827 were in the SB cohort and 150 in RB group. Median age and Charlson Comorbidity Index (CCI) were significantly higher in the SB cohort. 60% of SB patients had public insurance. Mortality was significantly higher in the SB group than the RB (38.2% vs 26%) (p = 0.004). In multivariate adjusted analysis (Table 1), residing in the SB (OR 1.803, 95% CI 1.121-2.901), age> 60 years old (OR 2.518, 95% CI 1.742-3.640) and critical COVID-19 infection (OR 14.559, 95% CI10.013-21.169) were significantly associated with mortality. Comparison of Hispanic (631) and Black (295) patients in the cohort showed higher median age and CCI in the Hispanic group (Table 3). There was no difference in mortality between the two cohorts.

**Table 1.** Univariate and multivariate analysis of the relationship between variables and mortality.

|  | Death, n (%) | Alive, n (%) | Univariate analysis |  | Multivariate analysis |  |
| --- | --- | --- | --- | --- | --- | --- |
|  | N=355 (36.3) | N=622 (63.7) | OR (95% CI) | p value | OR (95% CI) | p value |
| Community, South Bronx | 316 (89.0) | 511 (82.2) | 1.760 (1.191-2.602) | 0.004* | 1.803 (1.121-2.901) | 0.015* |
| Age, >60 years old | 252 (71.0) | 300 (48.2) | 2.626 (1.989-3.468) | <0.001* | 2.518 (1.742-3.640) | <0.001* |
| Sex, female | 152 (42.8) | 268 (43.1) | 0.989 (0.760-1.287) | 0.935 | 0.888 (0.642-1.227) | 0.471 |
| Race |  |  |  |  |  |  |
| Latinx | 239 (67.3) | 392 (63.0) | 1.209 (0.918-1.592) | 0.176 |  |  |
| Black | 96 (27.0) | 199 (32.0) | 0.788 (0.590-1.051) | 0.105 |  |  |
| Insurance |  |  |  |  |  |  |
| Uninsured | 47 (13.2) | 136 (21.9) | 0.545 (0.380-0.782) | 0.001* | 0.803 (0.496-1.302) | 0.374 |
| Private | 63 (17.7) | 162 (16.0) | 0.613 (0.442-0.849) | 0.003* | 0.759 (0.509-1.132) | 0.176 |
| Public | 245 (69.0) | 324 (52.1) | 2.049 (1.577-2.696) | 0.001* | 1.076 (0.902-2.133) | 0.056 |
| Household condition |  |  |  |  |  |  |
| Own/rent | 334 (94.1) | 580 (93.2) | 1.152 (0.671-1.978) | 0.608 |  |  |
| Long term nursing facility/prison | 11 (3.1) | 16 (2.6) | 1.211 (0.556-2.639) | 0.629 |  |  |
| Homeless | 5 (1.4) | 17 (2.7) | 0.508 (0.186-1.390) | 0.179 |  |  |
| Religious belief |  |  |  |  |  |  |
| Non-religious | 77 (21.7) | 143 (23.0) | 0.928 (0.678-1.270) | 0.640 |  |  |
| Christian | 248 (69.9) | 420 (67.5) | 1.115 (0.841-1.478) | 0.450 |  |  |
| Muslim | 11 (3.1) | 17 (3.7) | 1.138 (0.527-2.457) | 0.742 |  |  |
| Current smoker | 9 (2.5) | 28 (4.5) | 0.552 (0.257-1.183) | 0.121 |  |  |
| Alcoholism | 35 (9.0) | 76 (12.2) | 0.897 (0.467-1.980) | 0.311 |  |  |
| Illicit drug abuse | 14 (3.9) | 21 (3.4) | 1.098 (0.766-1.450) | 0.116 |  |  |
| Comorbidities |  |  |  |  |  |  |
| Obesity | 170 (47.9) | 280 (46.0) | 1.080 (0.831-1.402) | 0.566 |  |  |
| Asthma/COPD | 72 (20.3) | 109 (17.5) | 1.197 (0.860-1.667) | 0.286 |  |  |
| Hypertension | 160 (45.1) | 266 (42.9) | 1.098 (0.845-1.428) | 0.485 |  |  |
| Congestive heart failure | 32 (9.0) | 35 (5.6) | 1.662 (1.010-2.735) | 0.044* | 1.118 (0.598-2.091) | 0.727 |
| Diabetes mellitus | 185 (52.1) | 272 (43.7) | 1.400 (1.078-1.819) | 0.012* | 1.008 (0.800-1.536) | 0.538 |
| Chronic kidney disease | 65 (18.3) | 78 (12.5) | 1.563 (1.092-2.238) | 0.014* | 0.947 (0.593-1.512) | 0.819 |
| Cancer | 35 (9.9) | 37 (5.9) | 1.729 (1.068-2.800) | 0.024* | 1.463 (0.825-2.595) | 0.199 |
| Liver cirrhosis | 1 (0.3) | 7 (1.1) | 0.248 (0.030-2.205) | 0.270 |  |  |
| HIV | 9 (2.5) | 17 (2.7) | 0.926 (0.408-2.099) | 0.853 |  |  |
| COVID-19 severity, critical | 196 (55.2) | 52 (8.4) | 13.512 (9.497-19.226) | <0.001* | 14.559 (10.013-21.169) | <0.001* |

Variables that showed significant association with mortality entered the multiple logistic regression analysis to identify variables that are independently associated with high mortality.

**Table 2.**
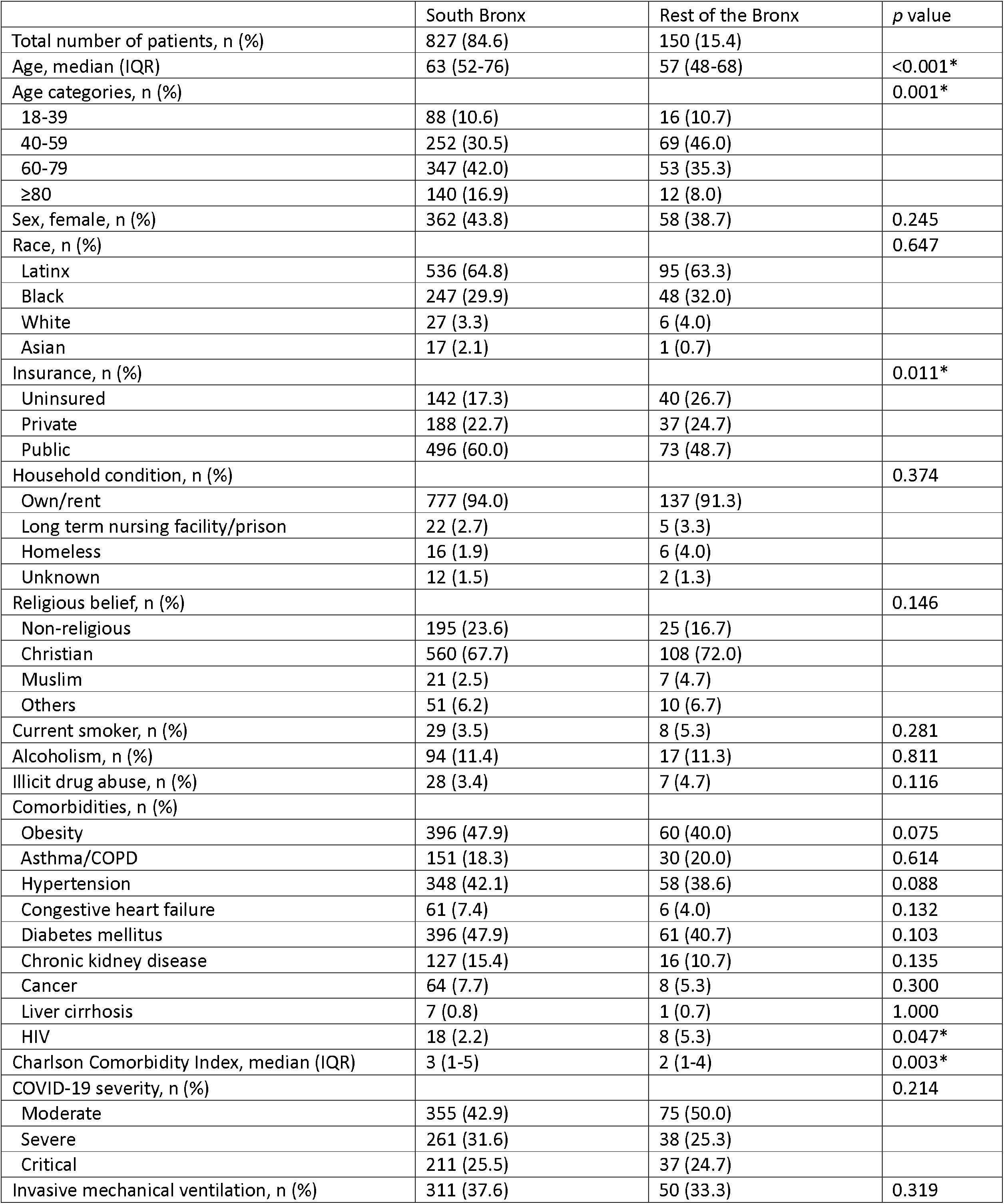
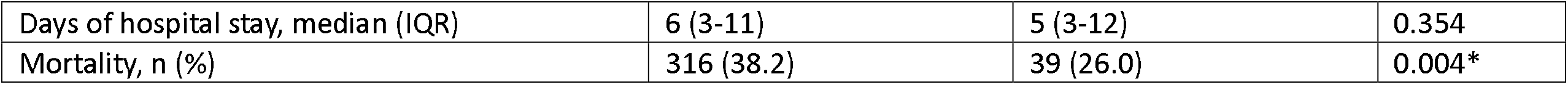
Characteristics of patients from south Bronx versus rest of the Bronx admitted for COVID-19 infection.

**Table 3.** characteristics of Latinx versus African-American patients admitted for COVID-19 infection.

|  | Latinx | Black | *p* |
| --- | --- | --- | --- |
| Total number of patients, n (%) | 631 (68.1) | 295 (31.9) |  |
| Age, median (IQR) | 64 (51-76) | 60 (59-72) | 0.008 |
| Age categories, n (%) |  |  | 0.167 |
| 18-39 | 61 (9.7) | 40 (13.6) |  |
| 40-59 | 204 (32.3) | 99 (33.6) |  |
| 60-79 | 259 (41.0) | 118 (40.0) |  |
| ≥80 | 107 (17.0) | 38 (12.9) |  |
| Sex, female, n (%) | 275 (43.6) | 120 (40.7) | 0.405 |
| Insurance, n (%) |  |  | 0.188 |
| Uninsured | 122 (19.3) | 55 (18.6) |  |
| Private | 133 (21.1) | 78 (26.4) |  |
| Public | 376 (59.6) | 162 (54.9) |  |
| Household condition, n (%) |  |  | 0.273 |
| Own/rent | 593 (94.6) | 274 (92.9) |  |
| Long term nursing facility/prison | 14 (2.2) | 11 (3.7) |  |
| Homeless | 16 (2.5) | 4 (1.4) |  |
| Unknown | 8 (1.3) | 6 (2.0) |  |
| Religious belief, n (%) |  |  | 0.505 |
| Non-religious | 142 (22.5) | 71 (24.1) |  |
| Christian | 431 (68.3) | 197 (66.8) |  |
| Muslim | 22 (3.5) | 6 (2.0) |  |
| Others | 36 (5.7) | 21 (7.1) |  |
| Current smoker, n (%) | 24 (3.8) | 10 (3.4) | 0.755 |
| Alcoholism, n (%) | 69 (10.9) | 38 (12.9) | 0.575 |
| Illicit drug abuse, n (%) | 22 (3.5) | 11 (3.7) | 0.896 |
| Comorbidities, n (%) |  |  |  |
| Obesity | 292 (46.4) | 136 (46.1) | 0.925 |
| Asthma/COPD | 107 (17.0) | 60 (20.3) | 0.212 |
| Hypertension | 261 (41.4) | 147 (49.8) | 0.016* |
| Congestive heart failure | 44 (7.0) | 17 (5.8) | 0.489 |
| Diabetes mellitus | 307 (48.7) | 125 (42.4) | 0.074 |
| Chronic kidney disease | 104 (16.5) | 33 (11.2) | 0.034* |
| Cancer | 47 (7.4) | 22 (7.5) | 0.996 |
| Liver cirrhosis | 7 (1.1) | 0 | 0.104 |
| HIV | 13 (2.1) | 11 (3.7) | 0.137 |
| Charlson Comorbidity Index, median (IQR) | 3 (1-5) | 3 (1-4) | 0.006* |
| COVID-19 severity, n (%) |  |  | 0.546 |
| Moderate | 273 (43.3) | 139 (47.1) |  |
| Severe | 195 (30.9) | 85 (28.8) |  |
| Critical | 163 (25.8) | 71 (24.1) |  |
| Invasive mechanical ventilation, n (%) | 239 (37.9) | 101 (34.1) | 0.284 |
| Days of hospital stay, median (IQR) | 6 (3-10) | 6 (3-11) | 0.799 |
| Mortality, n (%) | 239 (37.9) | 96 (32.5) | 0.116 |

## Discussion

Our study, while confirming the significant association of residence in a poor socioeconomic community with mortality, fails to demonstrate any association of race/ethnicity with poor outcome. The patients from the SB had higher number of comorbidities than RB, increasing their risk of mortality^6^. While poor housing, difficulty to socially distance and crowded neighborhoods increased their risk for infection^7^, it is suspected that the older patients delayed their hospitalization due to anxiety about healthcare exposure from the virus. Additionally, while Hispanic and Black patients from the Bronx had overall higher mortality when compared to the rest of NYC^5^, we did not observe any difference in mortality with respect to race/ethnicity in our cohort. Our results conform with the large National Cohort Study of 6 million US veterans that found no observed difference in 30-day mortality by race/ethnicity among those infected with COVID-19 in spite of the increased disease burden^8^.

The COVID-19 pandemic has differentially affected outcomes in the SB compared to the RB. Remarkably there were no racial/ethnic disparities among our hospitalized patients. It is imperative to address the social/economic factors contributing to healthcare access disparity to reduce the adverse impact of COVID-19 in these communities.

## Data Availability

No additional data to be added. All data referred to is within the manuscript.

